# Model for Ki-67 Proliferation Index Predicton, Trained End-to-End on Routine Diagnostic Data

**DOI:** 10.1101/2025.06.26.25330333

**Authors:** Adam Kukučka, Jan Obdržálek, Vít Musil, Rudolf Nenutil, Petr Holub, Tomáš Brázdil

## Abstract

For breast cancer, the Ki-67 index gives important information on the patient’s prognosis and may predict the response to therapy. However, semi-automated methods for Ki-67 index calculation are prone to intra-and inter-observer variability, while fully automated machine learning models based on nuclei segmentation, classification and counting require training on large datasets with precise annotations down to the level of individual nuclei, which are hard to obtain. We design a neural network that straightforwardly predicts the Ki-67 index from scans of H&DAB-stained tissue samples. The network is trained only on existing data from daily operations at Masaryk Memorial Cancer Institute, Brno. The image labels contain only the Ki-67 index without any tumour epithelium or nuclei annotations. We use a simple convolutional neural network, not biasing the network by incorporation of layers dedicated to epithelium or nuclei segmentation or classification. Our model’s predictions align with the state-of-the-art evaluation by pathologists using QuPath image analysis with manual tumour annotation. On a test set consisting of 1250 images, the model achieved the mean absolute error of 3.668 and Pearson’s correlation coefficient of 0.959 (p < 0.001). Surprisingly, despite using a simple architecture and very weak supervision, the model persuasively detects complex morphological structures such as tumour epithelium. The model also works on Whole Slide Image data, e.g. to detect the hotspot areas. Since our approach does not need any specifically labelled data or additional staining, it is cost-effective and allows easy domain adaptation.

## I. Introduction

Ki-67 is a nuclear protein tightly related to the cell cycle. It serves as a biological surfactant to disperse mitotic chromosomes [1]. A detectable level appears in the G1 phase, progressing through the S and G2 phases to mitosis. After mitosis is finished, the level drops suddenly, and the protein is not detectable until the cell stays out of cycle in G0 phase [2]. By detection and counting of Ki-67 positive and negative cells we can estimate the proliferation rate of a selected cell population.

This is of particular use in tumour pathology, where the fraction of Ki-67-positive tumour cells, the Ki-67 index (also called Ki-67 labelling index or proliferation index, abbreviated as PI), is often correlated with the clinical course of cancer. In particular, for the evaluation of breast cancer, the knowledge of the Ki-67 index is of the utmost importance, despite the still-ongoing debate concerning methodology, thresholding, and clinical validity [3]–[6]. The proliferation rate of breast cancer provides us with important information regarding patient prognosis, and under some circumstances, it also predicts the response of cancer to therapy [5], [7]. For this reason the evaluation of the Ki-67 index is a part of daily routine pathology practice when dealing with breast cancer samples.

Immunohistochemistry represents a dominant method of Ki-67 detection. It is based on the visualization of cell nuclei containing Ki-67 by DAB staining and counterstaining Ki-67 negative cells using hematoxylin; see Fig. 1a. This method is quite robust, can be standardized up to some extent, and consensual recommendations regarding the protocols are given [5], [6]. In contrast, quantifying the staining result represents a less definitively solved problem. Theoretically, manual counting done by simple clicking of positive and negative nuclei in the snapshot or whole slide image should represent a gold standard. In the pathology practice, this approach is too tedious and time-consuming if the tumour’s internal heterogeneity is to be representatively covered, as this requires counting in larger areas or more areas containing together at least 500 and preferably 1000 or more tumour cells, following the above-cited recommendations.

**Fig. 1.**
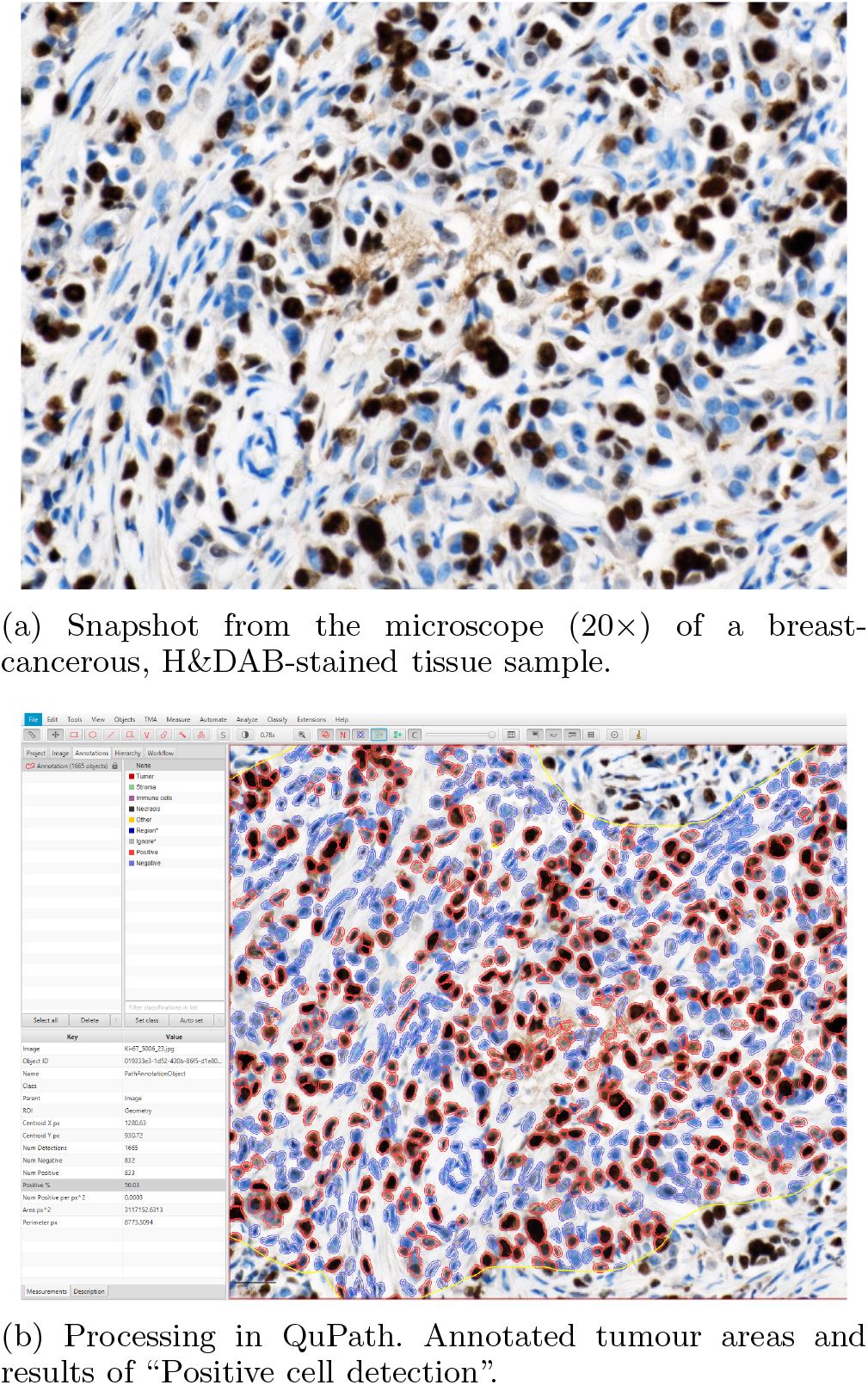
An example of clinical data is from one case at the Department of Pathology, Masaryk Memorial Cancer Institute, Brno. A Pathologist snapshots a hotspot and processes it in QuPath to get the Ki-67 index.

Unsurprisingly, a visual estimate by an experienced pathologist (often called “eyeballing”) initially became the prevalent method of Ki-67 evaluation in routine examinations. Due to the subjective nature of this approach, the reproducibility in the clinically most relevant category of grade 2 oestrogen receptor-positive carcinomas is not satisfactory, and also the results of exact counting need not be convincing as well [8].

(Semi-)automated platforms combining manual cancer delineation or machine learning with automated detection of positive and negative nuclei should represent a potential solution [6], [8]–[12]. The current standard, represented in the recommendation of International Ki-67 Breast Cancer Working Group [6], utilizes free software QuPath [13] trained to identify cells and discriminate between cancer and non-cancer cells. It counts the positive and negative cancer cells thresholded according to the mean value of nuclear DAB staining. Such an approach can be prone to stain variation (namely counterstain) and variable algorithm settings [14]. Also, the discrimination between cancer and non-cancer cells based on adaptive learning in detected objects may fail up to some extent in less common tissue patterns or in cancers exhibiting dissociated growth. In our study, we tested an alternative approach: a weakly supervised machine learning without any user intervention.

a) Paper contribution: We develop a method based on convolutional networks that gives a close estimate of the Ki-67 index on breast cancer tissue scans obtained from hematoxylin and DAB-stained tissue samples. More concretely, the input to our algorithm is an image covering an area of 1408×1024 pixels (resolution 0.486 *µ*m/px), and the output is the estimate of the Ki-67 index. This method does not require any manual intervention, and its predictions align with the evaluation by experienced pathologists.

We were able to develop our machine-learning model purely based on existing data from day-to-day operations at the Pathology Department, Masaryk Memorial Cancer Institute (MMCI), Brno. These data are being collected for diagnostic purposes and are neither intended nor carefully and precisely annotated to train machine learning models. Our main model is trained end-to-end using just the input images and their corresponding Ki-67 indices (as target values) estimated by pathologists.

Our method is thus a useful tool for fast routine diagnoses without going through full-scale WSI scanning. Note that our model also does not need any additional staining (in addition to hematoxylin and DAB staining) in order to identify the epithelium. This means our workflow is also time-and cost-effective and does not offset the time saved for the pathologist by extra expense and time spent in the lab.

From a technical point of view, we have developed a novel neural network structure specifically for predicting the Ki-67 index. The structure is biased toward this prediction by constraining the model to internally compute estimates of the number of positive and negative nuclei, as well as proportions of the epithelium tissue in small patches of the input image. Such a technique leads to better index prediction than a naive application of convolutional networks.

More importantly, our method allows us to extract an epithelium segmentation estimate that is used internally by the model to produce the index estimate assuming that also the numbers of positive and negative nuclei (for the whole input image) are used as target values during training. The user may use such insight to diagnose the model biases towards particular morphological structures and detect problems with input image parts.

### A. Related Work

The use of deep learning methods, typically using convolutional neural networks, for automating the task of Ki-67 index quantification is not new. Probably the first such application is that of [15]. Previously used computer methods included techniques such as thresholding, edge-based segmentation or clustering. For an overview, see [15]. Deep learning can be employed in multiple ways. One particular use is to automatically detect tumour regions and/or epithelium. For example [16] use an algorithm based on the ResNet to delineate cancerous regions and uses “deep residual learning” to count the nuclei statistics. Similarly, targeted PTM-NET [17] is based on the VGG-16 model. Recently, [18] detects a tumour in WSI. For a method using epithelial detection using deep learning, see e.g. [19].

Another approach is to utilize deep learning for Ki-67 nuclei segmentation, which can be subsequently used to compute the Ki-67 index, see e.g. [20], based on the well-known ResNet model. Particularly well-investigated are approaches based on the U-Net [21] model for biomedical image segmentation. For example [22] is a U-Net-based architecture which, unlike our model, is trained on individually annotated tumour nuclei, with distinctive colour markers for positive tumour cells, negative tumour cells, and the non-tumour cells/tissue. This requires having precisely annotated ground truth, which is costly and time-consuming. The performance of piNET compared to other techniques can be found in [23]. A related architecture, KiNet [24] has been developed for pancreatic datasets. Another improvement on the U-Net architecture is UV-Net [25], which seems to show slightly better results than both piNET and U-NET [21].

The efficiency of these methods has been recently investigated. [10] studies, on a sample of 90 pathologists, how much incorporating the UV-Net model improves the agreement, accuracy, and turnaround times for Ki-67 assessment. In a different direction, [23] compares manual counting against 5 digital analysis methods, including (semi-automatic) QuPath and (fully automatic) piNET.

All the above approaches rely on manually annotated datasets like cell or nuclei masks together with their labels, or masks of tumour or epithelial regions. This typically requires a lot of effort and time from trained pathologists, which is always in short supply.

## II. Material and methods

### A. Samples

We train and test our model on an image dataset of tissue samples of patients with breast cancer obtained during routine operations between March 2019 and March 2024 at MMCI. The samples were stained with hematoxylin and DAB as chromogen for the Ki-67 biomarker using standard Roche Ventana reagents and protocol with Roche antibody (30-9) in BenchMark Ultra autostainer.

A pathologist selects a hotspot, a sub-image of the WSI concentrating on the potentially cancerous tissue, and takes a snapshot with a microscopic camera; see Fig. 1a. The whole collection of snapshots was taken by three pathologists using Nikon microscopes Eclipse/Eclipse Ci-L equipped with cameras DS-L1/DS-Fi3. Both microscopes used Nikon objective Plan Apo 20×0.75. The images are stored in JPEG format with RGB channels, and their size varies between 1280×960 to 2880×2048 px depending on the camera model.

Snapshots were then processed in QuPath [13]. A pathologist first selects the tumour areas and then ran the “Positive cell detection” procedure. For each snapshot we thus also have a QuPath screenshot showing both the parameters for the “Positive cell detection” algorithm and the measurements obtained, including the percentage of Ki-67-positive nuclei. An example of an input snap-shot and its corresponding QuPath screenshot is shown in Fig. 1. We emphasize that all these steps are taken solely for the sake of the routine examination. We accessed the images and screenshots after they were created and we were not able to influence the process.

### B. Datasets

The images are downscaled to 1408×1024 pixels, preserving the aspect ratio by padding with white space if necessary. The resulting image resolution is approx 0.486 *µ*m/px. We extracted the Ki-67 index from the screenshots using an optical character recognition tool for approx. 90% of images and manually for the rest. Therefore, our dataset consists *𝒟* of pairs (*I, K*), where *I* is the rescaled snapshot image and *K* the Ki-67 index obtained from QuPath. We use this dataset to test the hypothesis that the Ki-67 index can be learned from routinely available data using an end-to-end machine-learning procedure.

However, the QuPath screenshots also contain measured counts of positive and negative nuclei, which we, due to their availability, extracted analogously to get an enhanced dataset 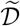 consisting of tuples (*I, K, P, N* ), where in addition, *P* denotes the number of positive and *N* the number of negative nuclei. We use this dataset to evaluate the relative contribution of the additional information present in the concrete numbers of positive and negative nuclei.

Firstly, we obtained 6040 cases collected between March 2021 and June 2023. The majority of cases are represented by a single snapshot; 368 cases have two snapshots, and 54 cases have three. In total, we have 6515 images. This set has been split randomly into training and preliminary testing sets in roughly 9/1 ratio, ensuring that all snapshots from a particular case are included in only one of the sets. The model itself is trained on the training set. Based on the preliminary testing set, we tuned the training hyperparameters. Before each training, we randomly cut off 10% from the training set to create a preliminary test set, which was used to monitor training. Once the model had been developed, we evaluated its performance on an independent final testing set of 1250 images of 1139 cases collected between July 2023 and March 2024. Reported results are with respect to this final testing set. The statistics for the training, preliminary testing, and final testing datasets are collected in Table I.

**TABLE 1.** Dataset statistics

|  | Cases | Images | Collection Date |
| --- | --- | --- | --- |
| Training | 5436 | 5870 | 03/2019–06/2023 |
| Preliminary testing | 604 | 645 |  |
| Final testing | 1139 | 1250 | 07/2023–03/2024 |

### C. Machine Learning Pipeline

We train a convolutional neural network-based model that predicts from a given 1408×1024 image the Ki-67 index. Even though the model is trained end-to-end to predict the index for the whole image, it internally estimates the epithelium and cell type distributions related to the predicted index value. Thus, it provides much deeper information related to the model prediction, which enhances its interpretability and trustworthiness.

The main idea is to make the prediction for much smaller patches to increase the granularity of the predictions. With smaller patches, we would get more local and finer information about where positive cells in tumour epithelium prevail. However, if the patches are too small, the network will have a very limited context, and reliable decisions about the cell types will not be feasible.

We decided to cut the input images into patches of 128×128 and embed these patches into slightly larger image tiles of 256×256 pixels (with white padding when there is no context on the borders) on which the network predictions are made. This turns out to be a reasonable balance between the locality requirement and the context availability.

Another problem arises as we have no ground truth labels for any smaller regions, which disallows us from training the network on individual tiles. Therefore, we decompose the whole input image into 11×8 non-overlapping patches, let the network predict from their tiles, and then aggregate. Unfortunately, the Ki-67 index is not decomposable in the sense that it is impossible to calculate the Ki-67 index for a union of disjoint regions from their individual Ki-67 indices.

To overcome this, we designed the network to predict the nuclei counts instead. Specifically, for an input tile *t ∈ I*, the network predicts the count of positive tumour nuclei 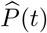 and the count of negative tumour nuclei 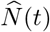. We then simply sum up these predicted counts across the patches and calculate the Ki-67 index 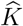 for the whole input image. Precisely, we denote

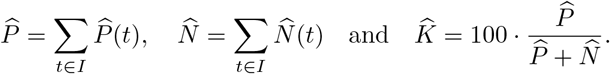

Note that the correctness of the above formula follows our inherent assumption that the model estimates 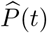 and 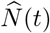 only from the patches embedded within the tiles, that is, from disjoint regions of the image.

We also write 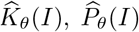 and 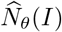 when we want to emphasize the dependency on a given image *I* and model weights *θ*. The forward pass of one input image is summarized in Fig. 2a.

**Fig. 2.**
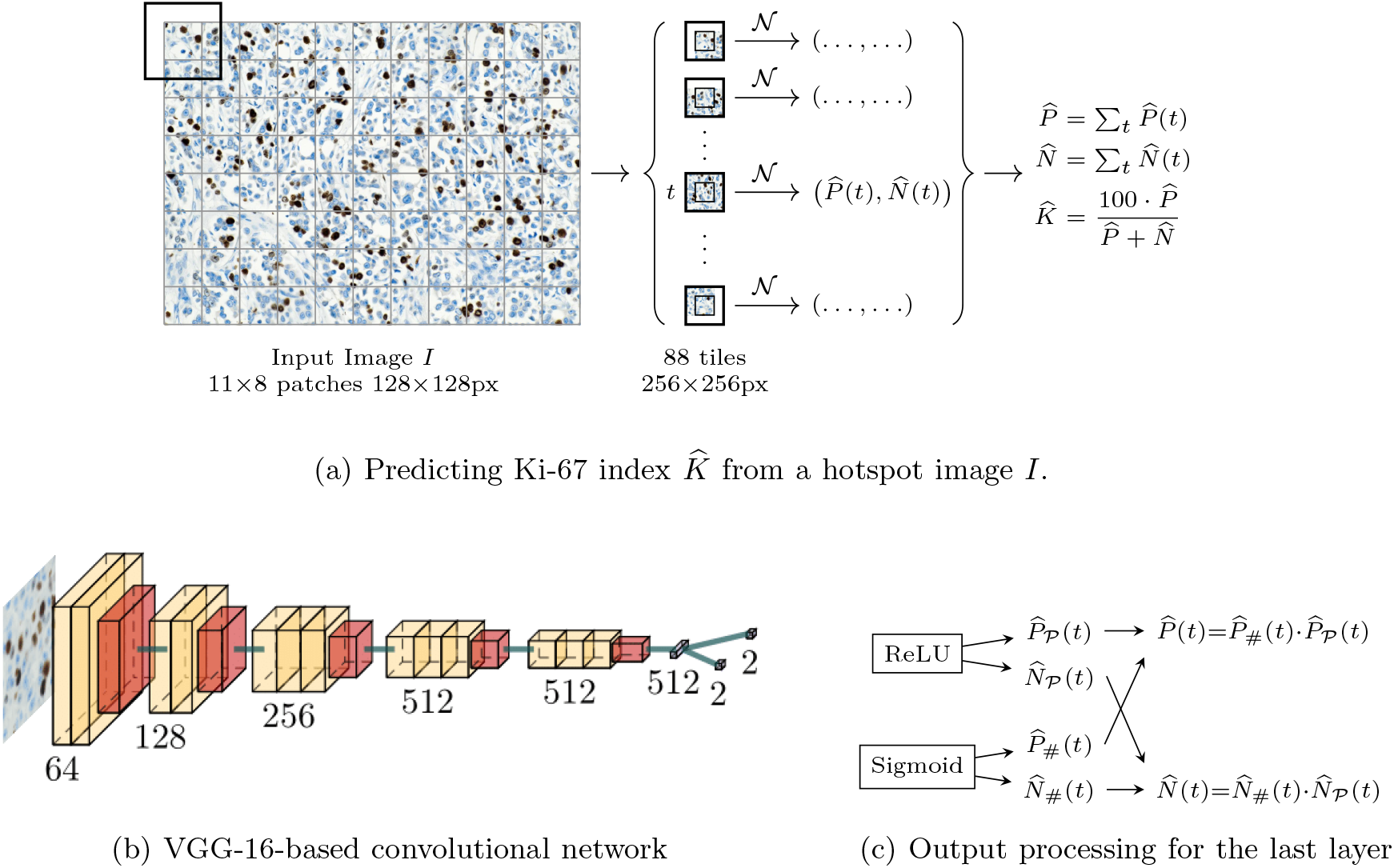
Model architecture. In (a) arrows labelled “*𝒩* “ refer to independent runs of the convolutional neural network, which predicts the number of positive 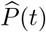 and negative 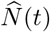 nuclei in a central patch from a slightly larger context tile *t*. The total counts of positive 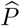, negative 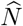 nuclei are aggregated, and the index 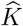 is computed.

### D. Model Training

In the simplest scenario, where only the ground truth Ki-67 index is available, we propose to train the model to minimize the empirical risk with a loss function measuring the squared error^1^ from the ground truth Ki-67 value, i.e.

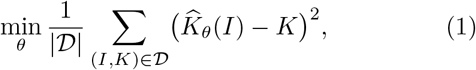

where *𝒟* is the training set. The trained model that aims to minimize (1) is denoted by *M*_K_.

Note that the Ki-67 index is a dimension-free variable, meaning that it does not carry any information about the magnitudes of the counts 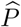 and 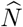, but only about their ratio. If we are interested in meaningful and explainable predictions also for the counts, one can empirically rescale the model outputs after the model is trained.

#### 1) Training with nuclei counts

Even though our main goal is to prove that a practically useful model can be trained only with the Ki-67 index as the target value, we also exploit the fact that the specific procedure used at MMCI also provides the values of the positive and negative nuclei counts (*P* and *N* ).

We train two models using this additional information and compare them with the main model trained only on the index. First, we consider training only with nuclei counts *P, N* as target values

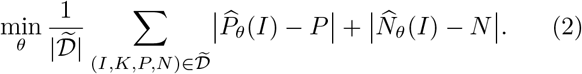

Note that the index is computed ex-post from using the counts. Second, we use both, the index *K* and the nuclei counts *P, N* as target values

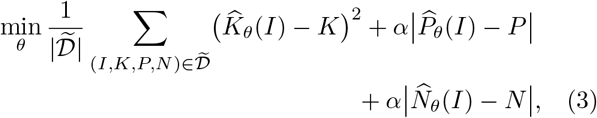

where *α* = 0.01 embodies the importance assigned to the nuclei count *P* (*N* ) predictions.

The models that are trained by optimizing (2) and (3) are denoted by *M*_PN_ and *M*_KPN_, respectively.

#### 2) Training process

All three models are trained using the same procedure. We use standard augmentations to enhance the training dataset: Random rotation, random flip, and random brightness/contrast increase/decrease by at most 20%.

We train the network for 20 epochs by stochastic gradient descent on mini-batches using Adam optimizer with a learning rate of 5 ·10^*−6*^. In our setup, the mini-batches must contain all tiles from the input images, and we process 4 images in a mini-batch.

### E. Neural Network Architecture

Our neural network, denoted *𝒩* in Fig. 2a, is based on the VGG-16 architecture [26]. Namely, we use VGG-16’s feature extractor, which consists of 13 convolutional and 5 max polling layers. We initialize the weights from the extractor pre-trained on the ImageNet (ILSVRC) 2012 data set [27]. Then, we follow with an adaptive average-pooling layer, effectively averaging each feature map to get a single feature vector of 512 dimensions. Finally, we build a nuclei count predictor from this feature vector.

The most trivial architecture would be a single 512×2 linear layer to get two numbers 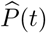 and 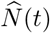. However, we decided to give the network a slightly larger expressibility by predicting the nuclei counts as a product of two interpretable numbers. Namely, we compute 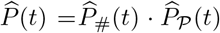, where 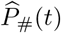 represents a total number of Ki-67 positive nuclei and 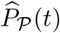 the proportion of positive nuclei which are in the epithelium. Analogously, 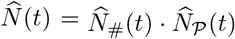, where 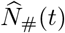 is the total number of Ki-67 negative nuclei and 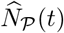 the proportion of negative nuclei contained in the epithelium. Therefore, we employ two512×2 linear layers, one followed by ReLU, to predict the absolute counts 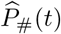 and 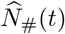, and the other, followed by the sigmoid function, to predict the proportions 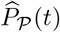 and 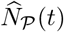 as shown in Fig. 2b, 2c.

This little enhancement will allow us to analyze and interpret the ability of the trained network to detect both nuclei types and epithelial structures. Note that there is no supervision on any of these four refined quantities, and their ground truth values are unknown to us. There is no guarantee that the network actually learns them; it is rather a designer’s apriori assumption of what they could be. Only posterior explainability analysis can test if this is true and hence allows us to interpret the network outputs in an understandable way.

### F. Model Inference

Recall that our trained network predicts the number of positive and negative nuclei in tumour epithelium given a 256×256-pixel tile. To predict the Ki-67 index on the testing hotspot image, we simply cut the image to non-overlapping 128×128-pixel patches embedded into 256×256-pixel tiles, obtain the predicted counts for all tiles, and then aggregate the counts to get the index, analogously as shown in Fig. 2a.

## III. Results

### A. Model Trained on Ki-67 Index

Our main concern is to decide whether a useful model can be trained using only the Ki-67 index as the target value. So, for this subsection, we consider a model *M*_K_ trained by solving the optimization problem (1).

We compare the output of our *M*_K_ model with the target values *K* from our dataset (obtained manually using QuPath; see Sec. II-B). Pearson’s correlation coefficient *R* = 0.959 (*p <* 0.001) indicates that our model predictions correspond to the values obtained using the semi-automatic procedure. A more detailed look at the absolute error (i.e., the absolute value of the difference between the model’s output and the target value) reveals that for more than half of the inputs, the absolute error is below 2.5 and for more than 3/4 of the inputs below 5. The mean absolute error is 3.668. The error distribution is shown in Fig. 6.

We also calculated the intraclass correlation coefficient (ICC) using the two-way mixed model with single measurements and absolute agreement, also known as ICC(3,1). The obtained value of 0.958 (95%-Confidence Interval 0.954–0.963) shows excellent reliability. Further statistics are reported in Fig. 3.

**Fig. 3.**
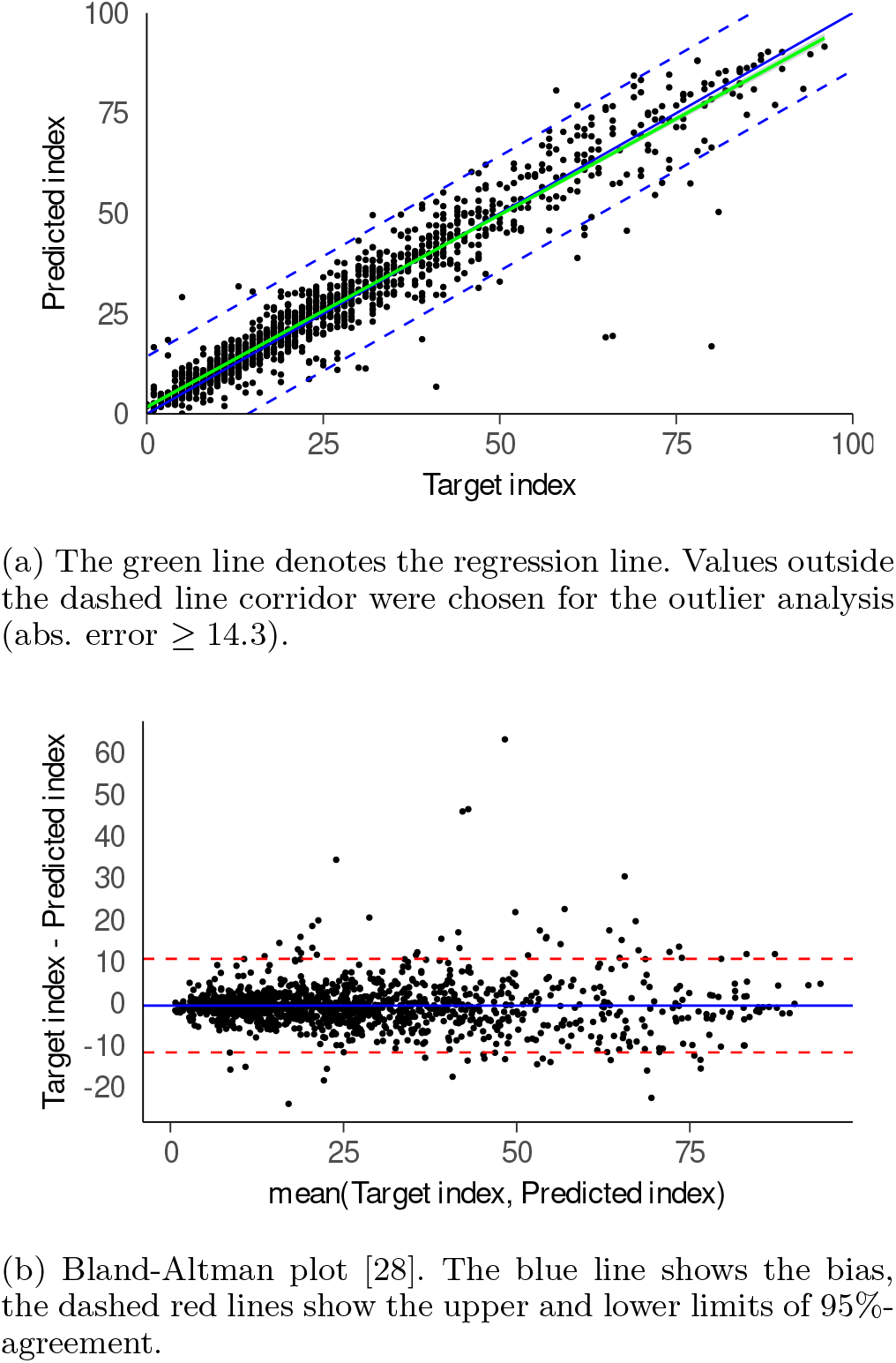
Results on the testing set of model *M*_K_, trained solely on the Ki-67 index.

#### 1) Outlier Analysis

The scatter plot Fig. 3a reveals several outliers where the model’s prediction strongly mismatches the target values from the test set. Recall that the training and testing data come from routine operations at MMCI; they were not sanitized in any way and can contain errors. Therefore, the outliers should be inspected manually to decide whether the error is on the side of the model or the dataset. For the manual inspection, we selected 30 images^2^ out of the 1250 images in the final testing set with the largest absolute error. These cases were analyzed by a pathologist with the following observations.

1) Data Error (8 of 30). The index in data was calculated from a different image than the prediction, the index in data was mistyped, or the input image contained non-cancerous epithelium. The model was neither trained nor intended for this situation. Also, the background of the DAB stained section caused model failure.

2) Primary Measurement Failure (8 of 30). The index in data was not properly measured by the pathologist (incomplete or wrong annotations, problems with hematoxylin counterstain not properly treated during QuPath use). All these cases could be curated, resulting in limited or no discrepancy.

3) Model Failure (14 of 30). Our model seems to fail in cases where it is not able to discriminate cancer from stroma, especially in the situation where the stroma is abundant, cellular, and the Ki-67 index of the stroma is much lower compared to cancer epithelium. In such cases, the predicted Ki-67 index is underestimated. Typical failures come in lobular cancer or in carcinomas with cellular lymphoid stroma, see Fig. 4.

**Fig. 4.**
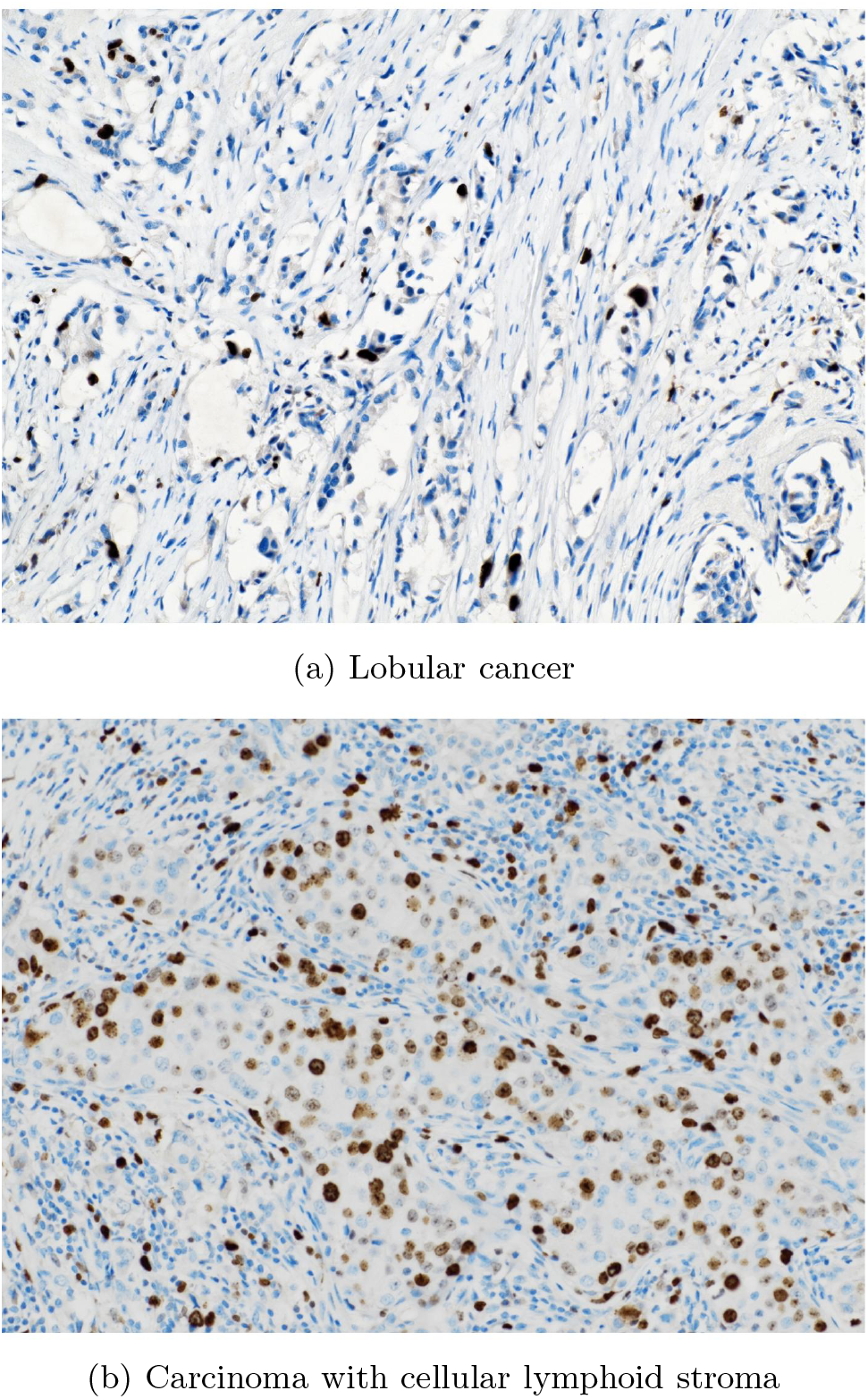
Examples of typical inputs, for which the predicted Ki-67 index is not accurate.

As we can see, only approximately 47% of outliers (2.4% of the test set) are due to model failure caused by images with ill-defined epithelium. The rest is caused by inadequate input images or data errors and are, in principle, curable. We can expect the training set to have a similar proportion of data errors and instrument failures. Nevertheless, we conclude that training the model on real-world, less-than-perfect ground truth still resulted in a well-performing model.

#### 2) “Eyeballing” Comparison

Before the advent of digital pathology and tools like QuPath, the traditional method to assess Ki-67 index (and similar metrics), has been ‘eyeballing’. Here a trained pathologist arrives at a rough figure of the index by estimating the percentage of positive nuclei in the specimen. The assessment is quite subjective but can be reasonably consistent for each individual pathologist. The principal advantage of eyeballing is its speed. We were therefore interested in how well our proposed method compares to the traditional eyeballing.

We selected 304 images randomly from the preliminary test set and let two pathologists estimate the Ki-67 index without actually counting the nuclei. We obtained three estimated indices for each hotspot. Two separate passes by pathologist A done several days apart, and a single pass for pathologist B. We compare all three indices predicted by pathologists and the index predicted by our model with the ground truth Ki-67 index obtained using QuPath. The statistics of the prediction errors are reported in Fig. 5. We can conclude that our model outperforms both trained pathologists regarding precision.

**Fig. 5.**
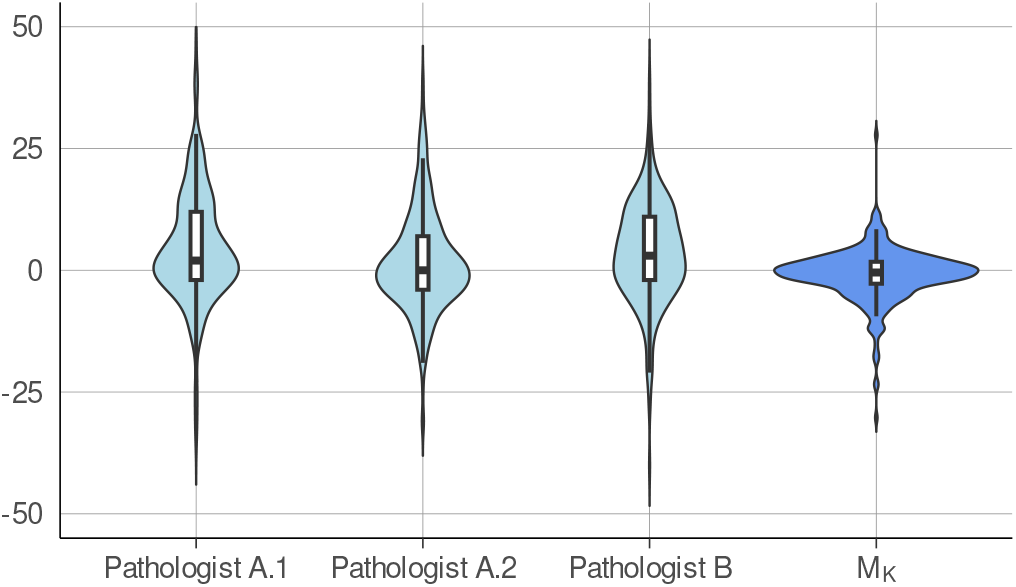
Distributions of the difference between the predicted Ki-67 index (either by a pathologist or our model *M*_*K*_) and the ground truth Ki-67 index obtained using QuPath on the preliminary test set.

**Fig. 6.**
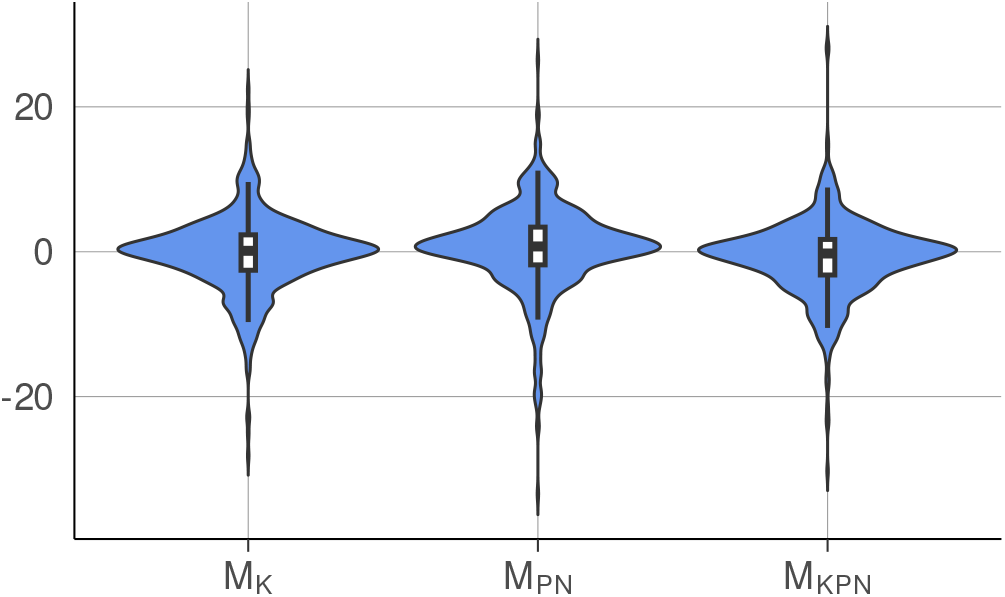
Distributions of the difference between the Ki-67 index predicted by each of the models and the ground truth Ki-67 index obtained using QuPath.

### B. Models Trained on Positive and Negative Nuclei Counts

As the dataset from MMCI contains not only the Ki-67 index but also the estimates of the counts of positive and negative nuclei, we have compared models using this additional information with the model trained solely to predict the index. We have trained two additional models:

- *M*_*PN*_ trained by solving the optimization problem (2) to predict solely the counts of the positive (*P*) and negative (*N*) nuclei where the Ki-67 index is computed ex-post using the predicted values of *P* and *N* .
- *M*_*KPN*_ trained by solving the optimization problem (3), where the model is trained to predict all three values, the index, and the counts of positive and negative nuclei.

We have evaluated these two models using the same approach as for *M*_K_; that is, we compared the output Ki-67 index with the index obtained using QuPath. The model *M*_PN_ achieves Pearson’s correlation coefficient *R* = 0.958 and the mean absolute error 4.191, while *M*_KPN_ has *R* = 0.962 with the mean absolute error 3.488. Detailed statistics are reported in Fig. 6.

Apparently, adding the prediction of the counts to the Ki-67 index predictor brings a small performance improvement. Predicting only the nuclei counts leads to worse results than the direct prediction of the index.

We have also evaluated the ability of the *M*_PN_ and *M*_KPN_ models to predict the counts *P* and *N* . We could also obtain the *P* and *N* counts from the *M*_K_ model but these would be typically scaled by a random linear factor, as the model had no information to train on. The count distributions are shown in Fig. 7. Surprisingly, the predictions of the counts are much less precise (especially for the negative nuclei counts) than the prediction of the index, which can be directly computed from the counts.

**Fig. 7.**
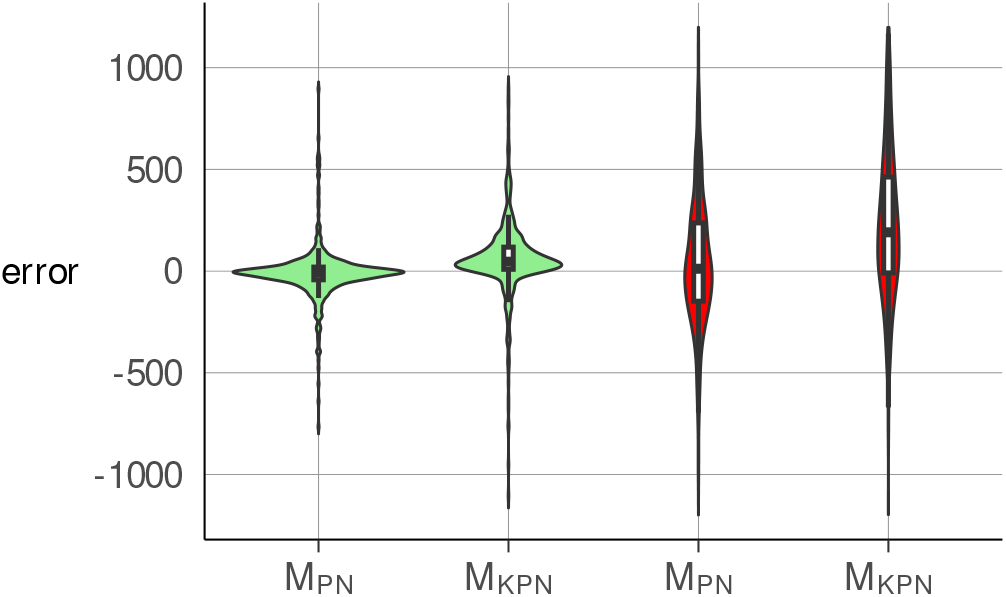
Distributions of the difference between predicted count of positive (green) and negative (red) nuclei by each of the models and the ground truth Ki-67 index obtained using QuPath.

## IV. Discussion

### A. Comparison to Other Methods

Direct comparison to competing methods on the same data is unfortunately not possible since the tools are either not accessible or the data inputs require different equipment and preprocessing. However, we can compare statistical results with methods reporting the same metrics. For the U-Net-based architecture piNET [22], its authors report the Pearson’s correlation coefficient to be 0.927 over all data and 0.966 over 256×256-sized hotspots. Our model *M*_K_ achieves comparable *R* = 0.959 without training on a finely annotated dataset ( [22] uses annotation down to single nuclei level).

### B. Model Inference on Whole Slide Images

In our model testing, we predicted the Ki-67 index for a hotspot image. Since our network is relatively small, running the predictions on the whole slide images is feasible. The aggregated Ki-67 index for the whole slide image may not be very informative, as the slide can also contain large proportions of non-tumour tissue, but it can be used as a guide to identify the hotspot areas.

To this end, we can partition the image into tiles/central patches (with the same 128px stride) and visualize the predictions as a heatmap, where the intensity of the color represents the predicted Ki-67 index. As the patch size is 128×128, such a heatmap would have a low resultion. To increase the resolution (and, simultaneously, to reduce the bias introduced by the choice of the tiling grid) we decided to increase the heatmap’s resolution by decreasing the stride to 16 pixels only. Any 16×16 region then overlaps with multiple central patches, and the color intensity of the resulting heatmap is therefore computed as an average over the index values for all contributing patches. An example of a Ki-67 heatmap computed in this way is shown in Fig. 8a. However, positivity heatmap alone does not contain enough information to identifity good hotspots. For the reasons explained in Sec. IV-D, we also need to be able to identify the distribution of epithelium, which we discuss next.

**Fig. 8.**
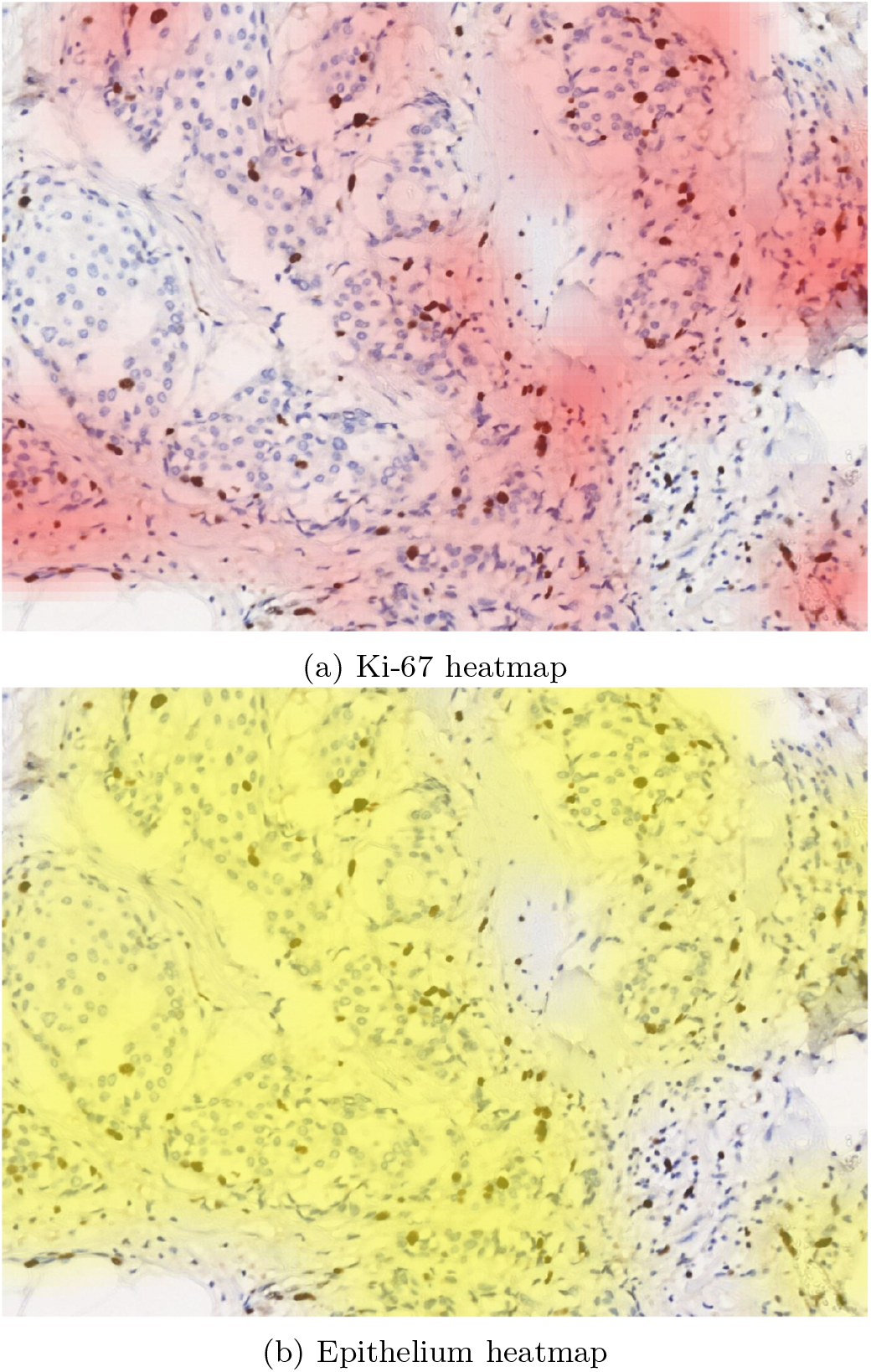
Heatmaps inferred by our model for the same input image. The alpha channel of the epithelium mask is reduced to half to better see underlying tissue.

### C. Model’s Ability to Detect Epithelium

By the design of the neural network, the model can predict not only the counts of positive and negative nuclei but also the proportion of these nuclei in the epithelium. This, in turn, allows us to predict the percentage of epithelium *Ê* (*t*) in a patch *t ∈ I* as

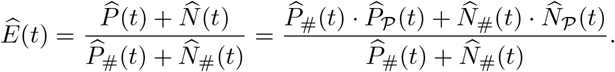

The values *Ê* (*t*) can be used to produce a heatmap of epithelium the same way as we did for the Ki-67 index, including the averaging of the overlapping patches strode by 16 pixels. An example of an epithelial heatmap is shown in Fig. 8b.

Visual inspection shows that the predictions of tumour epithelium are well aligned with reality. Typical examples can be seen in Fig. 9, showing both the model predictions and expert annotations of tumour epithelium. Moreover, since some of our data also contain QuPath annotations of tumour epithelium, we can assess the model’s capability to detect the epithelium quantitatively.

**Fig. 9.**
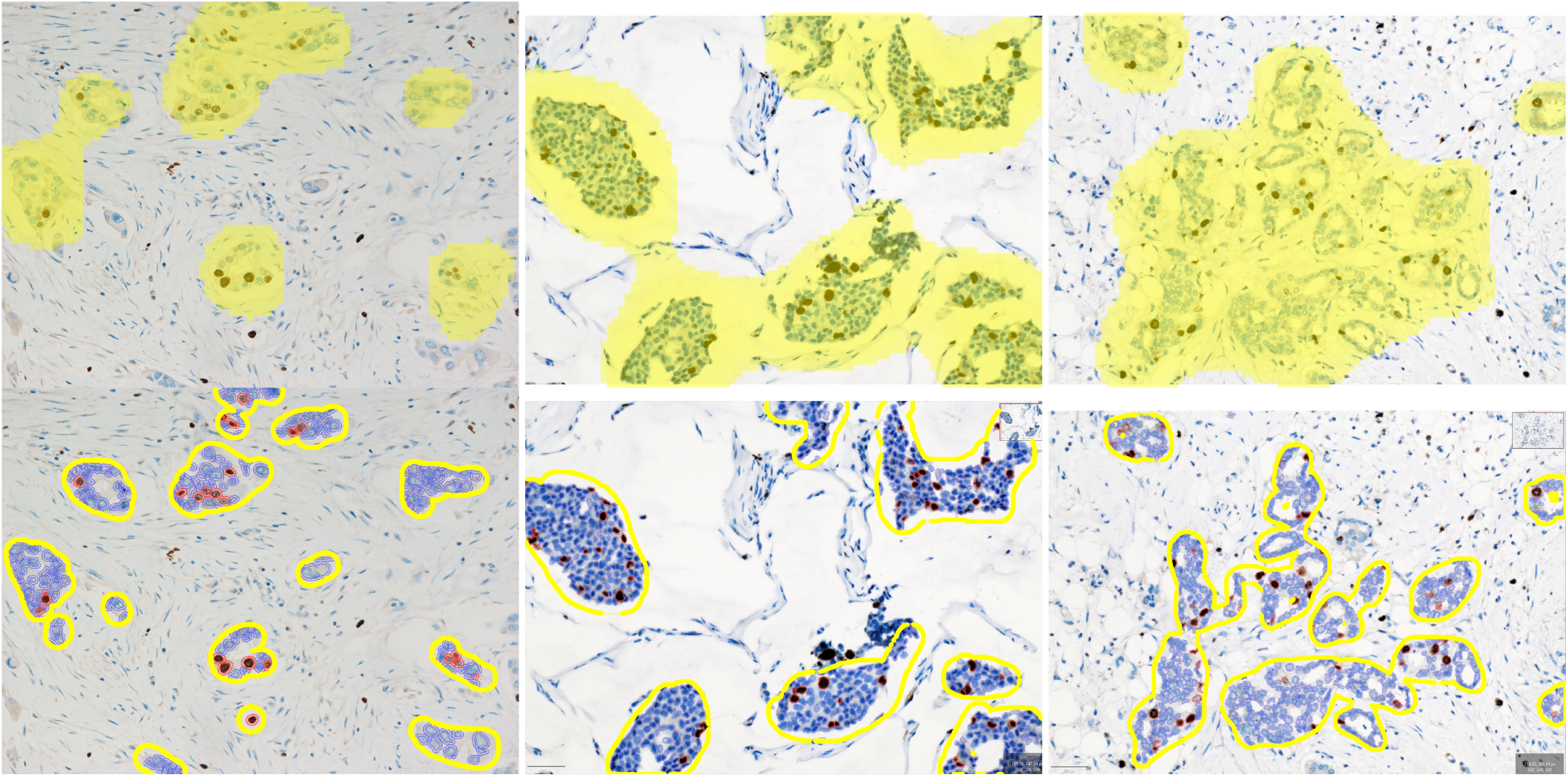
Three examples from the testing set with epithelium mask produced by our model on the top (yellow layer) and epithelium segments drawn by a pathologist in QuPath on the bottom (yellow countour).

In total, we collected ground truth epithelial masks for 220 images from the final testing set and evaluated a Jaccard similarity coefficient *J* (intersection over union). Next, our epithelial heatmaps attain arbitrary values between zero and one and have to be binarized on a certain threshold before the comparison. The maximal average Jaccard index is *J* = 0.57 *±* 0.04 for the threshold set to 0.66. Although the model was quite weakly supervised and was not trained to recognise epithelial structures, this result proves that it is indeed capable of detecting it, even if not perfectly.

However, the imperfection has two sources. One is the prediction errors, and the second is the design limitation. Indeed, even if the neural network perfectly tells the proportion of epithelium in all patches, the resulting heatmap lacks the expressivity to reproduce all various rich shapes, as it is constrained to averaging the patches and thresholding. To isolate the design error, we created the best possible heatmaps from ground truth epithelial masks by calculating the truth percentage of epithelium in each patch. Their average Jaccard index is *J* = 0.71 *±* 0.02 at the threshold 0.41, establishing the upper bound on the performance of any network. Therefore, the trained network achieves 0.57*/*0.71 *≈* 80%, which is impressive.

### D. Automatic Hotspot Detection on Whole Slide Images

To emulate the workflow of a pathologist, we can, given a whole slide image, select the hotspot on which to calculate the Ki-67 index. In principle, we are looking for region within a WSI, which has a high Ki-67 index. In our case, we seek for a 1408×1024 rectangle (consisitng of 11×8 pathes), but the process can be adjusted for different regions. We thus partition the input WSI into non-overlapping 128×128 patches and predict the values 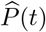 and 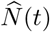 for each such patch (resp. the enclosing tile) *t*. The task is now reduced to finding an 11×8 rectangle of these patches, which would be our hotspot image *I*. The index value 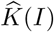 is then obtained by a simple numeric computation from the precomputed values 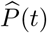 and 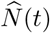 for all the patches included in *I*.

However, selecting the hotspot as the area with the highest Ki-67 index is too simplistic to work in practice. For instance, if a large region contains just a few nuclei and all of them are positive, the Ki-67 index will be high, but such a region is not suitable for the Ki-67 index measurement; see Fig. 10 for an example.

**Fig. 10.**
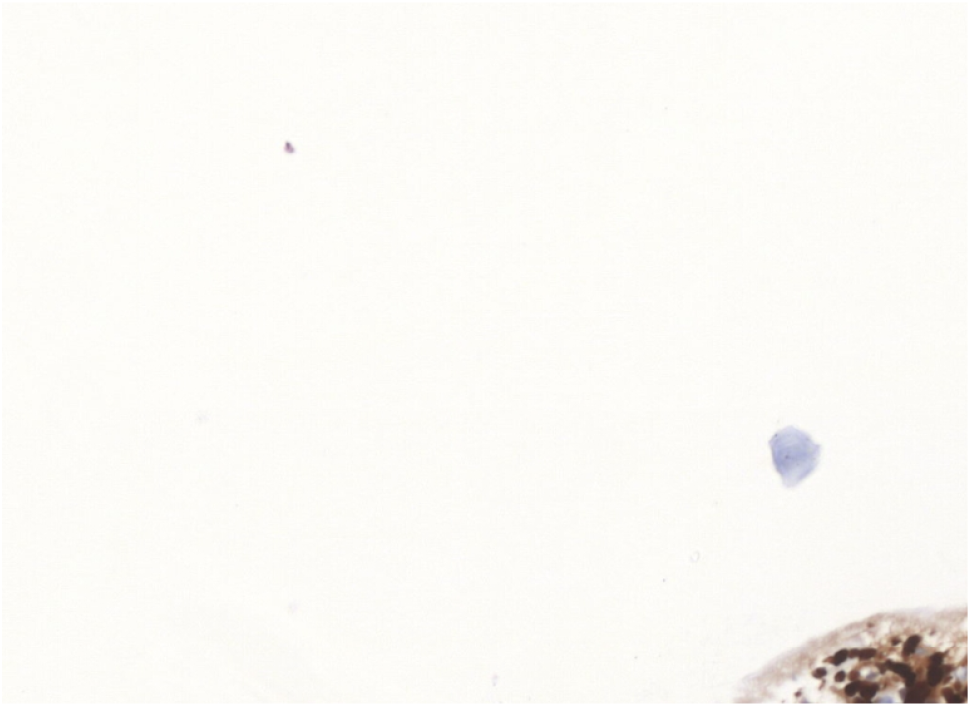
An example of an image with high Ki-67 index, which is not considered a hotspot, as it does not contain enough epithelium.

To counter this, we use the epithelium prediction from the previous section and consider only those 11×8 patch rectangles, which contain at least 25% of epithelium. This seems to work pretty well in practice, see Fig. 11 for a typical example, with the positivity heatmap produced as described in Sec. IV-B.

**Fig. 11.**
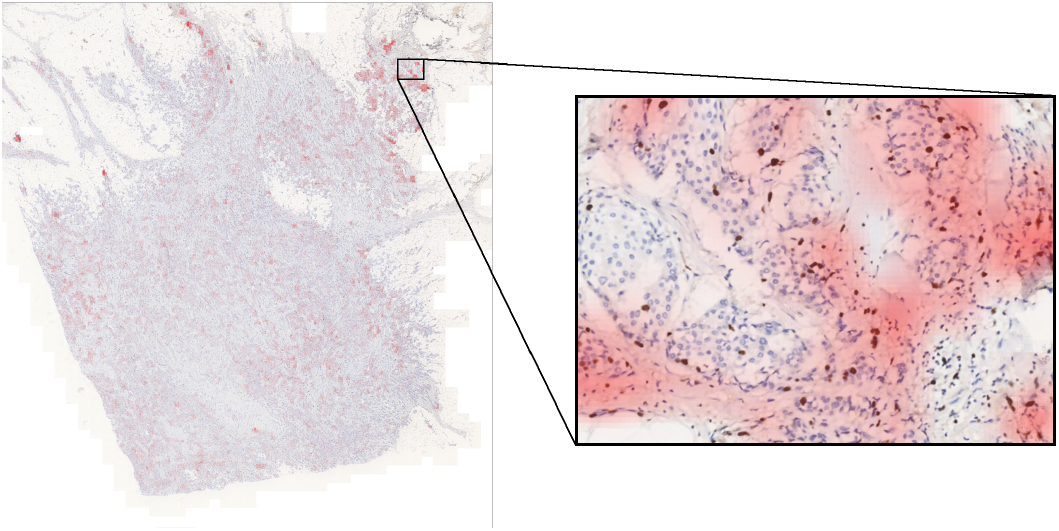
A WSI with an automatically selected hotspot. The intensity of the red colour shows the positivity heatmap.

## V. Conclusion

We have considered the problem of predicting the Ki-67 index on breast cancer tissue scans stained with hematoxylin and DAB. We concentrated on training a learning model that, for a given hotspot image, predicts the Ki-67 index, that is, the proportion of Ki-67 positive tumor cells in the epithelium tissue. We have shown that a high-quality model can be trained solely on data obtained from routine examinations at MMCI.

Concretely, we have trained our model to predict the Ki-67 index values obtained using a semi-automatic procedure utilizing QuPath. No additional detailed annotations of epithelium and/or Ki-67-positive cells are needed. This allows a simple domain adaptation for institutions doing routine Ki-67 index evaluation.

As for the technical contribution, we have developed a novel structure model (based on convolutional networks) tailored precisely to solve the prediction of the Ki-67 index. The model works better than a naive application of convolutional networks to the problem. Also, it allows direct inspection of its ability to correctly detect epithelium within which the proliferation index is being predicted.

As for future work, to allow sensible inspection of the epithelium segmentation capability, the model needs to predict not only the Ki-67 index, but also the counts of positive and negative nuclei. Our goal is to obtain such an interpretable model while using only the index as the target value during training. Another aim is to incorporate the model into the routine examination process and evaluate the domain adaptation capabilities in several hospitals.

## Data Availability

All data produced in the present study are available upon reasonable request to the authors.

## Footnotes

1 The choice of squared error is not essential. We have performed experiments which show that other loss functions, such as mean absolute error, perform comparably. This is also true for the loss functions used in (2) and (3) below.

2 This number was chosen so as not to overload our pathologist, and it corresponds to an absolute error of at least 14.3.

## Notes

This work has been supported by Czech Ministry of Health, (MMCI 00209805) and Czech Ministry of Education, Youth and Sports, (project BBMRI.cz, reg. no. LM2023033). AI infrastructure for the project was developed as a part of BioMedAI Center at Masaryk University, supported by the BioMedAI TWINNING project funded under EU Horizon Europe Programme, grant agreement no. 101079183.

### Competing Interest Statement

The authors have declared no competing interest.

### Author Declarations

Institutional Review Board of the Masaryk Memorial Cancer Institute, Brno, Czech Republic, gave ethical approval for this work.

## References

[1] S. Cuylen et al., “Ki-67 acts as a biological surfactant to disperse mitotic chromosomes,” Nature, vol. 535, no. 7611, pp. 308–312, Jul 2016.

[2] E. Endl and J. Gerdes, “The Ki-67 protein: Fascinating forms and an unknown function,” Experimental Cell Research, vol. 257, no. 2, pp. 231–237, 2000.

[3] M. G. Davey, S. O. Hynes, M. J. Kerin, N. Miller, and A. J. Lowery, “Ki-67 as a prognostic biomarker in invasive breast cancer,” Cancers, vol. 13, no. 17, 2021.

[4] A. Goldhirsch et al., “Personalizing the treatment of women with early breast cancer: highlights of the St Gallen international expert consensus on the primary therapy of early breast cancer 2013,” Annals of Oncology, vol. 24, no. 9, pp. 2206–2223, 2013.

[5] M. Dowsett et al., “Assessment of Ki67 in breast cancer: Recommendations from the International Ki67 in Breast Cancer Working Group,” JNCI: Journal of the National Cancer Institute, vol. 103, no. 22, pp. 1656–1664, 09 2011.

[6] T. O. Nielsen et al., “Assessment of Ki67 in breast cancer: Updated recommendations from the International Ki67 in Breast Cancer Working Group,” JNCI: Journal of the National Cancer Institute, vol. 113, no. 7, pp. 808–819, 12 2020.

[7] M. Gnant, N. Harbeck, and C. Thomssen, “St. Gallen/Vienna 2017: A brief summary of the consensus discussion about escalation and de-escalation of primary breast cancer treatment,” Breast Care (Basel), vol. 12, no. 2, pp. 102–107, 2017.

[8] Z. Varga et al., “How reliable is Ki-67 immunohistochemistry in grade 2 breast carcinomas? a QA study of the Swiss Working Group of breast- and gynecopathologists,” PLOS ONE, vol. 7, no. 5, pp. 1–12, 05 2012.

[9] B. Acs et al., “Ki67 reproducibility using digital image analysis: an inter-platform and inter-operator study,” Laboratory Investigation, vol. 99, no. 1, pp. 107–117, 2019.

[10] A. Dy et al., “AI improves accuracy, agreement and efficiency of pathologists for Ki67 assessments in breast cancer,” Scientific Reports, vol. 14, no. 1, p. 1283, Jan 2024.

[11] N. Abele et al., “Noninferiority of artificial intelligence–assisted analysis of Ki-67 and estrogen/progesterone receptor in breast cancer routine diagnostics,” Modern Pathology, vol. 36, no. 3, p. 100033, 2023.

[12] C. M. Focke, P. J. van Diest, and T. Decker, “St Gallen 2015 subtyping of luminal breast cancers: impact of different Ki67-based proliferation assessment methods,” Breast Cancer Research and Treatment, vol. 159, no. 2, pp. 257–263, Sep 2016.

[13] P. Bankhead et al., “QuPath: Open source software for digital pathology image analysis,” Scientific Reports, vol. 7, no. 1, p. 16878, 2017.

[14] P. Bankhead, “Developing image analysis methods for digital pathology,” The Journal of Pathology, vol. 257, no. 4, pp. 391–402, 2022.

[15] M. Saha, C. Chakraborty, I. Arun, R. Ahmed, and S. Chatterjee, “An advanced deep learning approach for Ki-67 stained hotspot detection and proliferation rate scoring for prognostic evaluation of breast cancer,” Scientific Reports, vol. 7, no. 1, p. 3213, Jun 2017.

[16] N. Xie et al., “Artificial intelligence scale-invariant feature transform algorithm-based system to improve the calculation accuracy of Ki-67 index in invasive breast cancer: a multicenter retrospective study,” Annals of Translational Medicine, vol. 10, no. 19, 2022.

[17] J. Joseph et al., “Proliferation Tumour Marker Network (PTM-NET) for the identification of tumour region in Ki67 stained breast cancer whole slide images,” Scientific Reports, vol. 9, no. 1, p. 12845, Sep 2019.

[18] Q. He et al., “Unsupervised domain adaptive tumor region recognition for Ki67 automated assisted quantification,” International Journal of Computer Assisted Radiology and Surgery, vol. 18, no. 4, pp. 629–640, Apr 2023.

[19] M. Valkonen et al., “Cytokeratin-supervised deep learning for automatic recognition of epithelial cells in breast cancers stained for ER, PR, and Ki-67,” IEEE Transactions on Medical Imaging, vol. 39, no. 2, pp. 534–542, 2020.

[20] Y. Liu et al., “Predict Ki-67 positive cells in H&E-stained images using deep learning independently from IHC-stained images,” Frontiers in Molecular Biosciences, vol. 7, 2020.

[21] O. Ronneberger, P. Fischer, and T. Brox, “U-net: Convolutional networks for biomedical image segmentation,” in Medical Image Computing and Computer-Assisted Intervention – MICCAI 2015, N. Navab, J. Hornegger, W. M. Wells, and A. F. Frangi, Eds. Cham: Springer International Publishing, 2015, pp. 234–241.

[22] R. S. Geread et al., “piNET – An automated proliferation index calculator framework for Ki67 breast cancer images,” Cancers, vol. 13, no. 1, 2021.

[23] M. Dawe et al., “Reliability and variability of Ki-67 digital image analysis methods for clinical diagnostics in breast cancer,” Laboratory Investigation, vol. 104, no. 5, p. 100341, 2024.

[24] F. Xing, T. C. Cornish, T. Bennett, D. Ghosh, and L. Yang, “Pixel-to-pixel learning with weak supervision for single-stage nucleus recognition in Ki67 images,” IEEE Transactions on Biomedical Engineering, vol. 66, no. 11, pp. 3088–3097, 2019.

[25] S. H. Mirjahanmardi et al., “Preserving dense features for Ki67 nuclei detection,” in Medical Imaging 2022: Digital and Computational Pathology, J. E. Tomaszewski, A. D. Ward, and R. M. L. M.D., Eds., vol. 12039, International Society for Optics and Photonics. SPIE, 2022, p. 120390Y.

[26] K. Simonyan and A. Zisserman, “Very deep convolutional networks for large-scale image recognition,” 3rd International Conference on Learning Representations, ICLR 2015 - Conference Track Proceedings, pp. 1–14, 2015.

[27] O. Russakovsky et al., “Imagenet large scale visual recognition challenge,” International Journal of Computer Vision (IJCV), vol. 115, no. 3, pp. 211–252, 2015.

[28] J. Martin Bland and D. Altman, “Statistical methods for assessing agreement between two methods of clinical measurement,” The Lancet, vol. 327, no. 8476, pp. 307–310, 1986, originally published as Volume 1, Issue 8476.

